# Anosmia in COVID-19 patients

**DOI:** 10.1101/2020.04.28.20083311

**Authors:** Daniel Hornuss, Berit Lange, Nils Schröter, Siegbert Rieg, Winfried V. Kern, Dirk Wagner

## Abstract

**Objectives:** Coronaviruses (CoVs) have a neuroinvasive propensity, and the frequently reported symptoms of smelling and taste dysfunction in many COVID-19 patients may be related to the respective capability of SARS-CoV2, the cause of the current pandemic. In this study we objectified and quantified the magnitude and underreporting of the smelling dysfunction caused by COVID-19 using a standardized test.

**Methods:** We conducted a prospective cross-sectional study comparing the proportion of anosmia using Sniffin-sticks in those reporting a loss of smell, in those who did not as well as in uninfected controls. The outcome of anosmic versus not anosmic patients were recorded during hospital stay and at day 15 on a six-category ordinal scale. The study was approved by the institutional review board, all participants consented to the study.

**Results:** 40% of 45 consecutive hospitalized COVID-19 patients and 0% of 45 uninfected controls consenting were diagnosed with anosmia. 44% of anosmic and 50% of hyposmic patients did not report having smelling problems. Anosmia or hyposmia was not predictive of a severe COVID-19 manifestation.

**Conclusions:** The majority of COVID-19 patients have an objective anosmia and hyposmia, which often occurs unnoticed. These symptoms may be related to the neuroinvasive propensity of SARS-COV-2 and the unusual presentation of COVID-19 disease manifestations.

## Introduction

Coronaviruses (CoVs) including SARS-CoV-2, the cause of the current pandemic of coronavirus disease 2019 (COVID-19), have a neuroinvasive propensity[1,2], with the olfactorial neurons being currently discussed as portal of entry for neuroinvasion [3] and a spread of CoVs after infection of neural cells from CNS to the periphery via a transneural route [1]. A relevant proportion of admitted COVID-19 patients report the disturbances of taste or smelling [4], without any other obvious cause like nasal obstruction or rhinorrhea, which may be related to this capability. In this study we objectified the magnitude of the smelling disorder caused by SARS-CoV-2.

## Methods

Burghart-Sniffin’-Sticks®, a widely used screening test for smelling disorders, was used; according to the manufacturers specifications anosmia, hyposmia and normosmia, were defined as correctly identifying 1–6, 7–10, and 11–12 odors, respectively [5,6]. We conducted a prospective cross-sectional study at the Medical Center - University of Freiburg, Germany in April 2020 comparing the proportion of anosmia in patients with positive PCR result for SARS-CoV-2 in nasopharyngeal swaps or sputum using Sniffin-sticks in those reporting a loss of smell, in those who did not as well as in uninfected patients and health care workers as controls. Patients younger than 18 years, with known smelling disorder or who did not consent to the study were excluded. Assuming a prevalence of anosmia of 5% in the uninfected control group [6], the sample size was calculated to test the null hypothesis that anosmia in COVID-19 is the same as in controls and to find a significant difference using the chi square test with a power of >90%. To compare clinical course (symptoms, laboratory values at the time of the Sniffin test) and outcome of anosmic versus not anosmic patients the worst outcomes during the hospital stay and at day 15 on a six-category ordinal scale (1. discharged; 2. hospitalized, not requiring supplemental oxygen; 3. hospitalized, requiring supplemental oxygen; 4. hospitalized, on (NIV) or high flow oxygen devices; 5. hospitalized, on invasive mechanical ventilation (IMV) or extracorporal membrane oxygenation (ECMO); 6. death) were recorded.

The study was approved by the University hospital ethical committee (No. 184/20), written informed consent was obtained from all participants in the study. Study protocol and data set are available from the corresponding author upon request.

## Results

We tested 45 consecutive hospitalized COVID-19 patients and 45 uninfected controls (age (median years ± STD) 56 ± 16.9 and 54 ± 18.3, respectively) consenting to the study (table). The controls correctly identified a median of 11 out of 12 odors of Sniffin’ Sticks, none was anosmic, 12/45 (27%, 95%CI 14–41%, age (median years ± STD) 63 ± 19.6) were hyposmic, and 33 (73%, 95%CI 58–85%, age (median years ± STD) 49 ± 10.2) were normosmic. A higher percentage (18/45, 40%) of COVID-19 patients were diagnosed with anosmia (p< 0,001; table). COVID-19 patients smell on average 4 sticks less than uninfected controls (figure). The Sniffin’ Stick test was more sensitive in detecting anosmia in comparison to self-reporting or taking a medical history: 44% of anosmic and 50% of hyposmic patients did not report having smelling problems. The clinical picture, laboratory test results, and outcome at day 15 or by counting the worst outcome during the hospital stay defined by a rating on a 6-point ordinal scale was similar in patients with and without anosmia or hyposmia (table).

## Discussion

Hyposmia and anosmia are symptoms often noticed by COVID-19 patients [4]. Using a quantitative and objective test almost half of the patients were anosmic, and another 40 % hyposmic. Still in our cohort only 49% of patients reported a smelling dysfunction, which is more than in a recently published survey [4], but significantly less in comparison to what was diagnosed by the Sniffin’ test. E.g. the magnitude of the olfactory dysfunction in COVID-patients is underreported with more than 80% of COVID-19 patients having hyposmia or anosmia, in comparison to the uninfected controls, where no participant was anosmic and 27% hyposmic. In a large German community sample (n = 7267) a similar low percentage of anosmia (5%) has been reported using the same test; however the 12-stick-test is not able to distinguish properly between hyposmia and normosmia [6], thus the high percentage of 44 % of our COVID-19 patients with hyposmia needs to be interpreted carefully. Patients were not tested after having been discharged, but telephone interviews even with patients with mild to moderate COVID-19 showed that not all patients had returned to normal smelling 15 days after start of first symptoms although no other symptoms persisted.

Olfactorial neurons are discussed as portal of entry for neuroinvasion of CoVs which may be transferred to the Central Nervous System (CNS) via a synapse-connected route [2]. It is unclear whether olfactory sensory neurons are directly involved in the pathogenesis of smelling loss in COVID-19, since they have not been shown to coexpress Transmembrane protease, serine 2 (TMPRSS2) and Angiotensin Converting Enzyme 2 (ACE2), which are central for entry of SARS-CoV-2 into human cells [7]. In fact, the detection of the expression of these proteins in coexpression in olfactory sustentacular cells [7] does not preclude the direct involvement of olfacory sensory neurons in COVID-19 associated smelling loss. In a mouse model infected with another human coronavirus, e.g. HCoV-OC43, viral antigen is detected only in the olfactory bulb after 3 and in the whole brain tissue after 7 days [8]. Given the wide distribution of the Angiotensin Converting Enzyme 2 (ACE2)-receptor in the brain [9], the observation that HCoV are able to induce direct neuronal injury within brainstem cardiorespiratory centers in experimental animal models [2] and the increasing evidence that SARS-CoV 2 is also causing neurological complications, the clinical presentation of COVID-19 patients with deterioration at around one week and the acute respiratory failure may be related to the neuroinvasive potential of SARS-CoV-2 [10].

In conclusion all COVID-19 patients should be interviewed and – if possible tested - for olfactory disorders that occur often in COVID-19. Some patients have presented solely with this symptom, e.g. primary physicians and otolaryngologist need to be aware of this putative presentation. Our study shows that anosmia and hyposmia often occurs unnoticed in COVID-19 patients, and that for those patients an objective and quantifiable test is required. Anosmia is not a predictor of a severe COVID-19 manifestation.

## Data Availability

Study protocol and data set: Available from Prof. Dr. D. Wagner upon legitimate request.

## Funding

This work was supported by the Berta-Ottenstein-Programme for Clinician Scientists, Faculty of Medicine, University of Freiburg, to NS.

## Acknowledgements

We sincerely thank Kamilla Szabo, M.D. and Annegrit Decker, M.D. (Department of Internal Medicine II, Medical Center – University of Freiburg, Germany) for taking care of the patients during the study allowing DH to test the participants.

## Reproducible Research Statement

Study protocol and data set: Available from Prof. Dr. D. Wagner (e-mail).

## Disclosure

The authors report no disclosures relevant to the manuscript

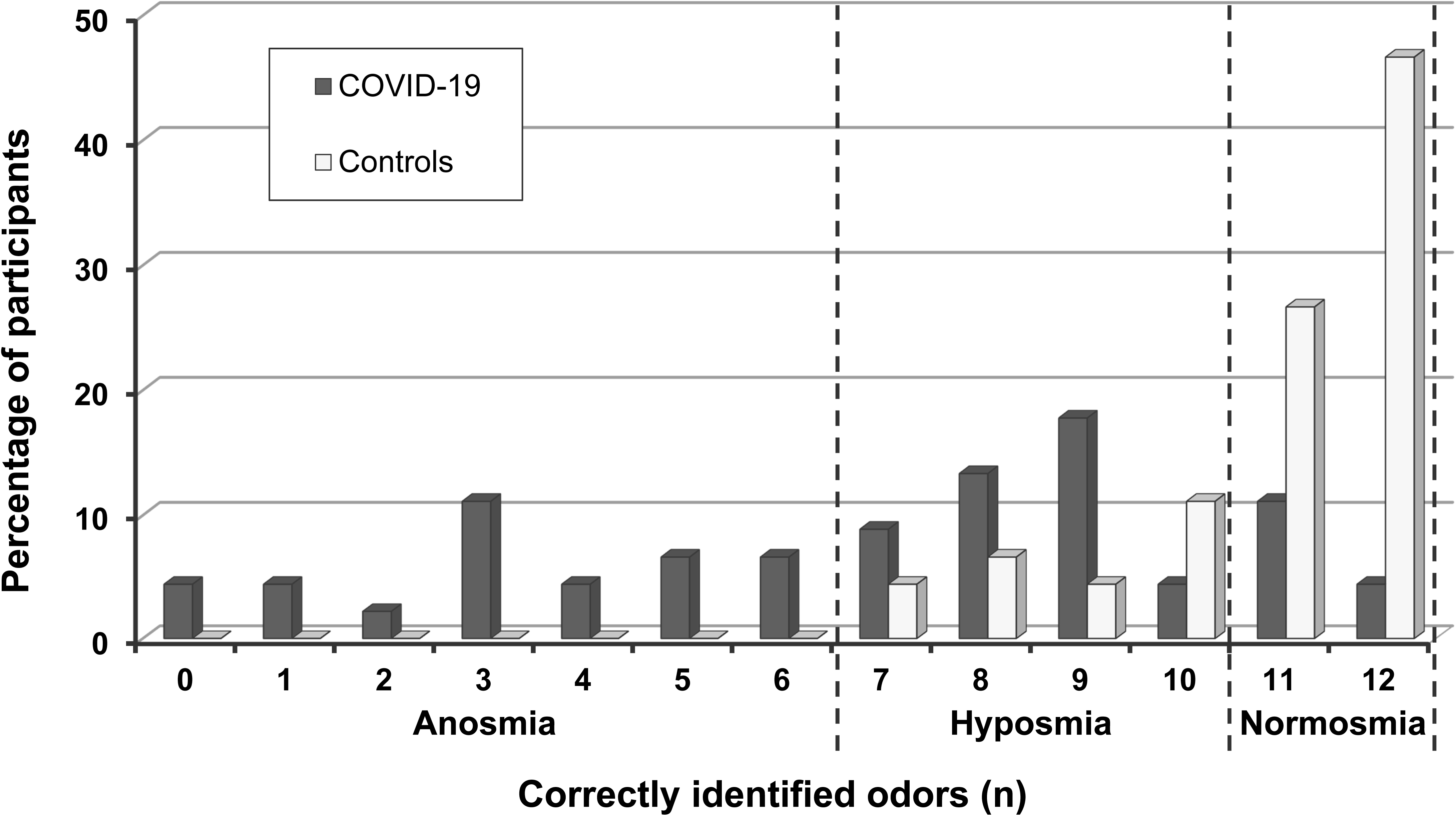
Figure: Number of correctly identified odors. The Sniffin test uses 12 different odors; the percentage of participants (y-axes; COVID-19 patients black bars; controls white bars) correctly identifying the respective number (n, x-axes) of odors is shown. The dashed lines define the cutoff between anosmia, hyposmia, and normosmia according to the test guidelines [5].

---

**Table:** Characteristics and results of COVID-19 patients.

**Table: Characteristics and results of COVID-19 patients.**
|  |  | all | with anosmia * | with hyposmia | with normosmia |
| --- | --- | --- | --- | --- | --- |
| All (n, %, 95 CI) |  | 45 | 18 (40%; 25-55%) | 20 (44%, 29-60%) | 7 (15%, 5-25%) |
| Sex (n) | female | 20 | 8 | 8 | 4 |
|  | male | 25 | 10 | 12 | 3 |
| Age (median ± STD) | Years | 56 ± 16.9 | 56.5 ± 17.9 | 58 ± 17.5 | 49 ± 10.2 |
| Symptoms (n, %) | Temp ≥ 38,5°C | 35 (78%) | 12 (34%) | 16 (46%) | 7 (20%) |
|  | Cough | 28 (62%) | 14 (50%) | 10 (36%) | 4 (14%) |
|  | Headache | 10 (22%) | 6 (60%) | 4 (40%) | 0 |
|  | Shortness of Breath | 20 (44%) | 13 (65%) | 5 (25%) | 2 (10%) |
| Laboratory values (mean ± STD) § | Leukocytes (Ths/μl] | 5.6 ± 2.8 | 6.3 ± 3.3 | 5.5 ± 1.9 | 4.3 ± 3.8 |
|  | Lymphocytes (Ths/μl] | 1.1 ± 0.7 | 0.8 ± 0.6 | 1.1 ± 0.8 | 1.4 ± 0.8 |
|  | CRP (mg/l) | 49 ± 50 | 60 ± 42 | 46 ± 49 | 19 ± 75 |
|  | PCT (ng/ml) | 0.12 ± 1.3 | 0.13 ± 0.1 | 0.17 ± 2.0 | 0.1 ± 0.1 |
|  | IL6 (pg/ml) | 26 ± 44 | 16 ± 34 | 27 ± 51 | 47 ± 44 |
|  | LDH (U/l) | 284 ± 181 | 294 ± 258 | 271 ± 148 | 249 ± 75 |
| Sniffin Test (median) | correctly identified odors # | 8 | 3 | 9 | 11 |
| Report of impaired smelling (n, %, 95CI) ¶ | yes | 22 (49%) | 10 (45%, 24-68%) | 10 (45%, 24-68%) | 2 (10%, 1-29%) |
|  | no | 23 (51%) | 8 (34%, 16-57%) | 10 (43%, 23-65%) | 5 (21%, 7-43%) |
| Time course (days; median ± STD) | first symptom to first positive PCR result | 2 ± 4.2 | 2 ± 4.1 | 1.5 ± 3.7 | 2 ± 4 |
|  | first symptom to reported impaired smelling | 5 ± 3.04 | 6.5 ± 3 | 2.5 ± 2.7 | 5.5 ± 2.5 |
|  | first symptoms to Sniffin test | 10 ± 5.1 | 11 ± 4 | 8.5 ± 4.5 | 12 ± 7.5 |
|  | first positive PCR result to Sniffin test | 4 ± 4.6 | 3.5 ± 3.9 | 4 ± 4.4 | 5 ± 6.3 |
| Clinical course during hospital stay (n, %) ± | Discharged | 23 (51%) | 11 (48%) | 9 (39%) | 3 (13%) |
|  | No oxygen | 27 (68%) | 8 (30%) | 13 (48%) | 6 (22%) |
|  | Supplemental oxygen | 15 (33%) | 7 (47%) | 7 (47%) | 1 (7%) |
|  | NIV or high flow oxygen | 2 (4%) | 2 (100%) | 0 | 0 |
|  | IMV or ECMO | 0 | 0 | 0 | 0 |
|  | Death | 1 (2%) | 1 (100%) | 0 | 0 |
| Outcome at day 15 (n, %) † | Discharged | 17 (41%) | 7 (41%) | 6 (35%) | 4 (24%) |
|  | No oxygen | 14 (32%) | 3 (21%) | 8 (57%) | 3 (21%) |
|  | Supplemental oxygen | 9 (22%) | 7 (78%) | 2 (22%) | 0 |
|  | NIV or high flow oxygen | 0 | 0 | 0 | 0 |
|  | IMV or ECMO | 0 | 0 | 0 | 0 |
|  | Death | 1 (2%) | 1 (100%) | 0 | 0 |

## Notes

### Competing Interest Statement

The authors have declared no competing interest.

